# Comparison of the spatiotemporal characteristics of the COVID-19 and SARS outbreaks in mainland China

**DOI:** 10.1101/2020.03.23.20034058

**Authors:** Xi Zhang, Hua-Xiang Rao, Yuwan Wu, Yubei Huang, Hongji Dai

## Abstract

**Background:** Both coronavirus disease 2019 (COVID-19) and severe acute respiratory syndrome (SARS) are caused by coronaviruses and have infected people in China and worldwide. We aimed to investigate whether COVID-19 and SARS exhibited similar spatial and temporal features at the provincial level in mainland China.

**Methods:** The number of people infected by COVID-19 and SARS were extracted from daily briefings on newly confirmed cases during the epidemics, as of Mar. 4, 2020 and Aug. 3, 2003, respectively. We depicted the spatiotemporal patterns of the COVID-19 and SARS epidemics using spatial statistics such as Moran’s *I* and the local indicators of spatial association (LISA).

**Results:** Compared to SARS, COVID-19 had a higher incidence. We identified 3 clusters (predominantly located in south-central China, highest RR=135.08) for COVID-19 and 4 clusters (mainly in Northern China, highest RR=423.51) for SARS. Fewer secondary clusters were identified after the “Wuhan lockdown”. The LISA cluster map detected a significantly high-low (Hubei) and low-high spatial clustering (Anhui, Hunan, and Jiangxi, in Central China) for COVID-19. Two significant high-high (Beijing and Tianjin) and low-high (Hebei) clusters were detected for SARS, although the global Moran’s *I* value was not significant.

**Conclusions:** The different spatiotemporal clustering patterns between COVID-19 and SARS could point to changes in social and demographic factors, local government containment strategies or differences in transmission mechanisms between these coronaviruses.

## Introduction

Since the World Health Organization (WHO) declared the outbreak of coronavirus disease 2019 (COVID-19) a Public Health Emergency of International Concern (PHEIC) on Jan. 30 2019, this emerging infectious disease quickly spread in China and to other countries beyond China. As of Mar 4, the total number of confirmed cases of COVID-19 climbed to approximately 80,000, with more than 3,000 reported deaths. Approximately 670,000 people had been identified as close contacts of infected patients, and 32,870 people had been under medical observation or quarantine in China [1].

Compared to the severe acute respiratory syndrome (SARS) outbreak in 2003, which was also caused by a similar coronavirus, COVID-19 has been much more transmissible and rapidly spread from a single city to the entire country in just 30 days [2]. The estimated basic reproductive numbers (R0s) for COVID-19 and SARS were approximately 3.1 [3] and 2.7 [4], respectively. The transmission mechanisms of COVID-19 are currently poorly understood, although this disease is considered to be one of the most widespread and destructive infectious diseases. There is a need for a more integrated investigation and coordinated international response to the outbreak. Spatiotemporal analyses, which integrate spatial and time-series analyses, can provide additional information on the persistence of patterns over time and illuminate any unusual patterns.

Therefore, in this study, by collecting the daily numbers of newly confirmed COVID-19 and SARS cases during the two epidemics, we aimed to determine the spatial behavior and temporal features of the COVID-19 spread in mainland China and compared then with the respective features from the SARS epidemic using spatiotemporal analysis.

## Methods

### Data source

The present study included incident cases of COVID-19 and SARS in 31 provinces (provincial-level regions on the Chinese mainland). Incident cases infected by COVID-19 were extracted from the daily briefings on novel coronavirus cases from Jan. 20 to Mar. 4, 2020, provided on the official website of the National Health Commission of the People’s Republic of China [5]. We confirmed the daily total numbers of reported cases with the surveillance data provided by the WHO [6].

Incident cases of SARS were extracted from daily situation reports for mainland China from Apr. 21 to Aug. 3, 2003, which were posted by China.org.cn (in Chinese) and were also provided by the National Health Commission. We confirmed the daily total numbers of reported cases of SARS with the cumulative numbers of reported cases provided by the WHO [7]. The SARS data were left-truncated when day-by-day data before Apr. 21, 2002 were not available for each province.

### Case definition

Cases of COVID-19 included diagnosed cases confirmed by at least one of the following three methods: isolation of COVID-19 virus, at least two positive results for COVID-19 virus by real-time reverse-transcription polymerase chain reaction (RT-PCR) assay or a genetic sequence that matches COVID-19 virus [8]. A clinically diagnosed case was defined as a suspected case with imaging features of pneumonia, which has only been only applicable in Hubei Province since Feb. 12, 2020 [9].

Cases of SARS were defined in accordance with the “National Case Definition of Infectious Atypical Pneumonia (SARS) in China, 2003,” which was updated by the National Health Commission on Apr. 23, 2003. Criteria for probable and suspected SARS included a) travel to a SARS epidemic area in the 2 weeks before onset of symptoms or close contact with a probable SARS patient; b) fever of > 38°C; c) chest X-ray abnormalities; d) normal or decreased leukocyte count; and no response to treatment with antimicrobial drugs [10].

### Statistical analyses

We used ArcGIS software v10.2.2 (ESRI Inc., Redlands, CA, USA) to depict the spatial distribution and perform global and local spatial autocorrelation analyses. We used Kulldorff’s space-time scan statistical analysis to detect the space-time clusters of SARS and COVID-19 and to verify whether the geographic clustering was caused by random variation. Considering the relatively low incidence rate, we used the discrete Poisson probability model as the scanning statistical model. In Kulldorff’s space-time scanning, the radius of the population coverage was used, and the maximum spatial scanning area was set to cover 10% of the risk population. The maximum temporal scanning window was set to cover 50% of the total research time. The scan window was increased gradually from 0 to the maximum, and the log-likelihood ratios (LLRs) were calculated for each window. The window with the maximum likelihood was defined as the most likely cluster area. Other clusters with statistically significant LLRs were defined as the secondary potential clusters. The LLR *P*-value was estimated through 99,999 Monte Carlo simulations. A *P*-value < 0.05 indicated a significantly high risk inside of the scan window and a potential high-risk cluster of the disease. The relative risk (RR) of the disease in each cluster was calculated to evaluate the risk of SARS and COVID in the detected cluster areas.

The results of spatiotemporal scans are sensitive to various parameters, such as the maximum spatial and temporal cluster sizes. Thus, the selection of the maximum radius of the spatial scanning window and the maximum length of the temporal scanning window were very important. In this study, we selected the maximum spatial cluster sizes as 10% and 30% of the total population at risk and the maximum temporal cluster sizes as 50% of the total study period. Based on the optimal spatiotemporal parameters, retrospective space-time scanning analysis was applied to identify the geographic areas and time periods of potential clusters with significantly higher COVID-19 and SARS incidence than those of nearby areas.

The spatial autocorrelation analysis was conducted by using Open GeoDa software v1.2.0 (GeoDa Center for Geospatial Analysis and Computation, Arizona State University, AZ, USA). To identify the spatial clustering of the COVID-19 and SARS incidence at the provincial level, we used row standardized first-order contiguity Rook neighbors as the criterion for identifying neighbors, as described in [11]. We calculated Moran’s *I* value and the local indicators of spatial association (LISA) statistic to analyze the global and local clusters as well as spatial outliers. There were four categories of spatial patterns in the LISA map. The high-high and low-low locations (positive local spatial autocorrelation) were typically referred to as spatial clusters, while the high-low and low-high locations (negative local spatial autocorrelation) were termed spatial outliers. A cluster was computed as such when the value at a location (either high or low) was more similar to its neighbors than would be the case under spatial randomness. The high-high locations referred to hot spot areas where the risk of case spreading was higher than average, whereas the low-low locations referred to cool spot areas where the risk of case spread was lower than average.

Considering the stringent measure of quarantining in Wuhan (Hubei) and neighboring cities introduced on Jan. 23, 2020, we further conducted subgroup analyses by dividing the COVID-19 data into two stages: stage 1 (Jan. 20 to Feb. 6, 2020, quarantine date plus a 14-day incubation period) and stage 2 (Feb. 7 to Mar. 4, 2020). We also performed spatiotemporal clustering analysis for COVID-19 by excluding cases in Hubei.

## Results

### Temporal trends and patterns

As of Mar. 4, 31 provinces (100% of mainland China) reported 80,409 COVID-19 cases, with the number of incident cases ranging from 1 to 15,153 per day. The average incidence rate was 5.76 infections per 100,000 persons (range: 0.03-114.02) during the selected period of the COVID-19 epidemic. Outside of the Hubei Province epicenter, Beijing and Shanghai were among the first case-reported provinces for COVID-19 on Jan. 20, 2019. Compared with COVID-19, SARS had a less widely influential area but a longer epidemic duration, and only 24 provinces (77% of mainland China) reported 3,571 SARS cases as of Aug. 3, 2003, with an average incidence rate of 0.41 per 100,000 (range: 0.00-16.72). (**Fig. 1**).

**Figure 1.**
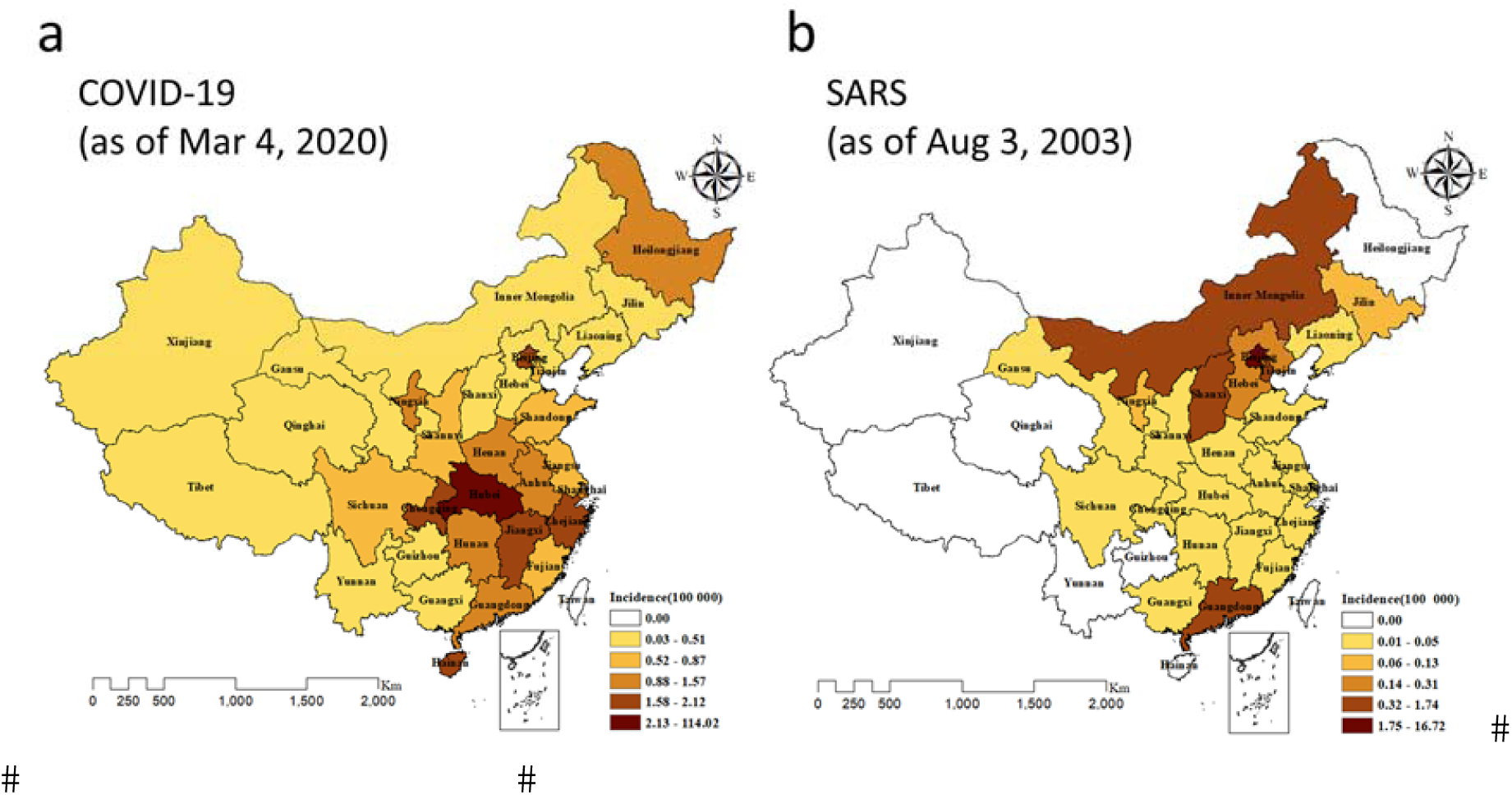
Spatial distribution of COVID-19 (a) and SARS (b) in mainland China.

To illustrate the spread of the two diseases nationally, we plotted the temporal changes in COVID-19 and SARS in 31 provinces in mainland China (**Fig. 2**, ordered by administrative area code). In most provinces except Hubei, the rate of increase in the number of cases for COVID-19 was fast for the first two weeks and reached a peak at the end of January. On the other hand, the incidence trend for SARS was mostly flat, except in Beijing, Tianjin, Hebei, Shanxi and Inner Mongolia. Notably, compared to SARS, there was an obvious increasing trend for COVID-19 in terms of the number of new cases in 12 provinces, such as Heilongjiang, Shanghai, Jiangsu, Zhejiang, Anhui, Jiangxi, Shandong, Henan, Hubei, Hunan, Chongqing and Sichuan. On the other hand, several provinces in western China, such as Guangxi, Yunnan, Shaanxi, Gansu, Qinghai, Ningxia, Xinjiang and Tibet, had a much lower prevalence for both COVID-19 and SARS.

**Figure 2.**
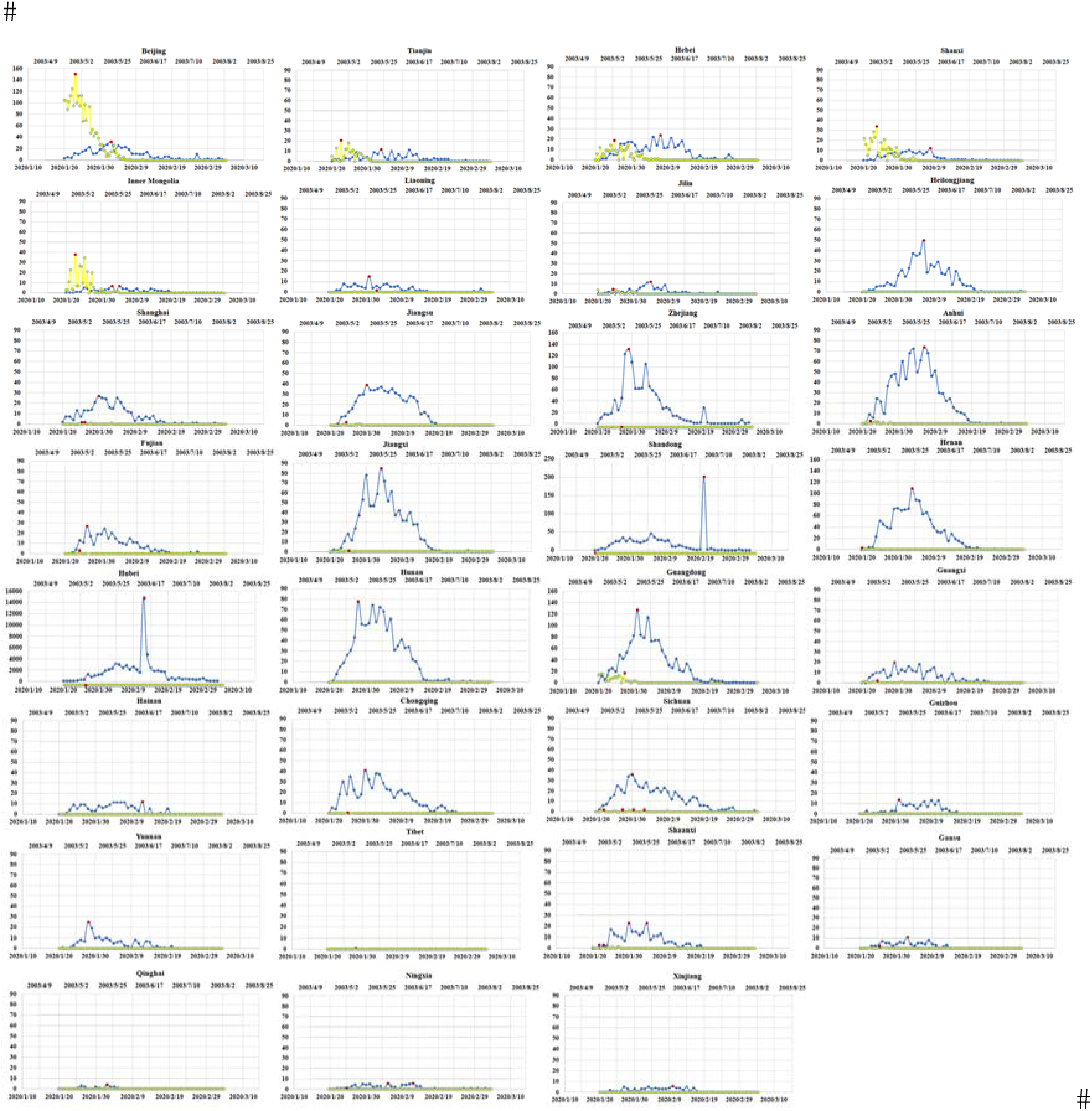
Temporal changes in the incidence of COVID-19 and SARS in 31 provinces in mainland China. The blue line indicates temporal changes for COVID-19 from Jan 20 to Mar 4, 2020. The yellow line indicates temporal changes in SARS from Apr 21 to Aug 3, 2003. The red dot indicates the peak number of incident cases.

### Identification of spatiotemporal clusters

Through spatiotemporal clustering analysis, we identified 4 high-risk clusters for COVID-19 within 4 cluster time frames (**Fig. 3a**). The most likely cluster was the epicenter, Hubei, with an RR of 135 compared with the neighboring provinces and the longest high-risk period of 22 days (*P* < 0.001). Two significant secondary clusters were identified in Zhejiang (from Jan. 28 to Jan. 30, 2020, *P* < 0.001) and Shandong (in Feb. 20, 2020, *P* < 0.001), with similar RRs of 1.64 and 1.56, respectively. Another possible cluster was identified in Jiangxi (from Feb. 3 to Feb. 4, 2020, *P* = 0.982).

**Figure 3.**
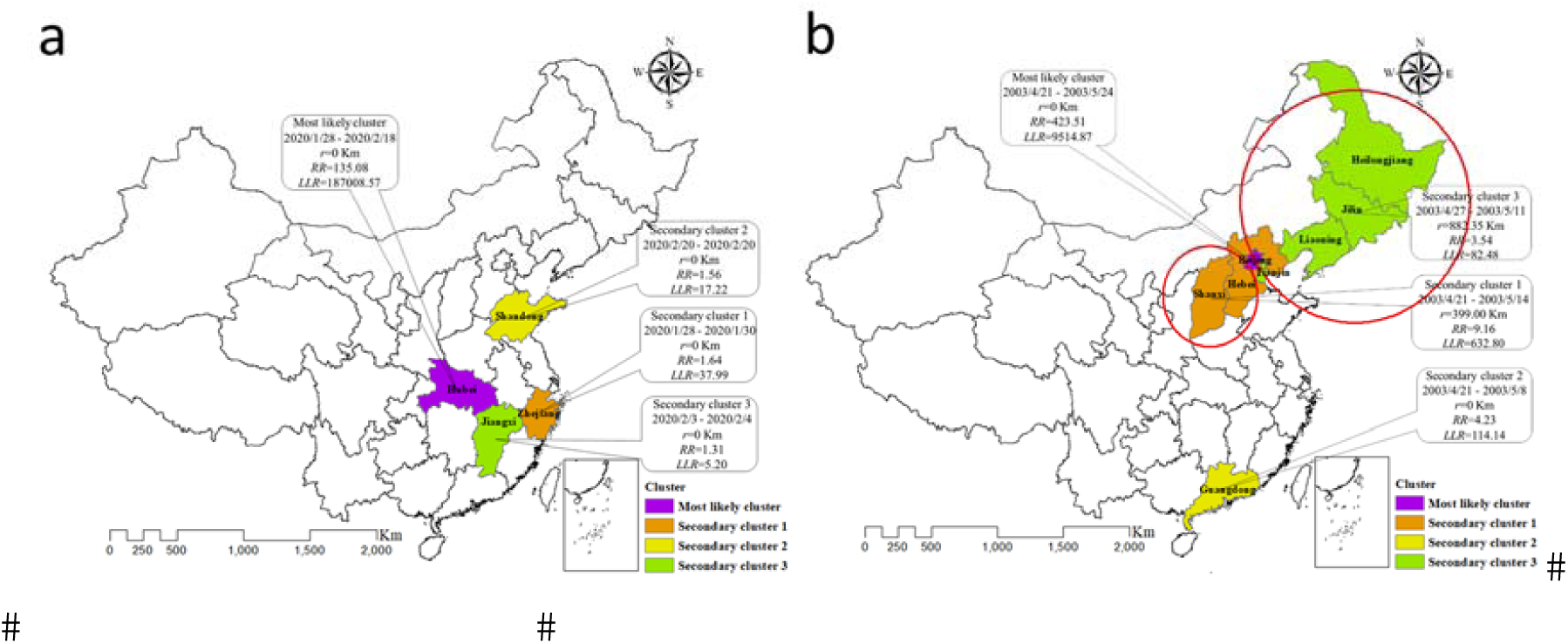
Comparison of spatiotemporal clustering of COVID-19 (a) and SARS (b).

When considering the measure of quarantine in Hubei, the RR of 223 in stage 2 (from Feb. 7 to Mar. 4, 2020) was largely increased compared to the RR of 69 in stage 1 (Jan. 20 to Feb. 6, 2020) (**Supplementary Fig. 1**). There were different spatial behaviors and temporal features between the two stages. When excluding cases in Hubei, the high-risk clusters were centered on the areas around Hubei and in Beijing, Shanghai, and Heilongjiang in stage 1, whereas the high-risk clusters were only restricted within the neighborhood areas of Hubei in stage 2. Moreover, the RRs in both stages were significantly decreased for the most likely cluster, with RRs of 3.56 in stage 1 and 5.31 in stage 2 (**Supplementary Fig. 2**).

Different from COVID-19, the most likely cluster of SARS was centered on Beijing (**Fig. 3b**), lasting from Apr. 21 to May. 24, 2003, with the highest RR of 423 and a longest period of 34 days (*P* < 0.001). Three significant secondary clusters were identified in Shanxi and Hebei (from Apr. 21 to May. 14, 2003, *P* < 0.001), Guangdong (from Apr. 21 to May. 8, 2003, *P* < 0.001), and provinces of Jilin, Liaoning, Heilongjiang and Tianjin (from Apr. 27 to May. 11, 2003, *P* < 0.001), respectively.

### Spatial autocorrelation

The global Moran’s *I* values for COVID-19 and SARS were -0.022 and 0.073, respectively (both *P* > 0.05), which indicated that the case distribution may have been due to chance rather than global autocorrelation (**Fig. 4**).

**Figure 4.**
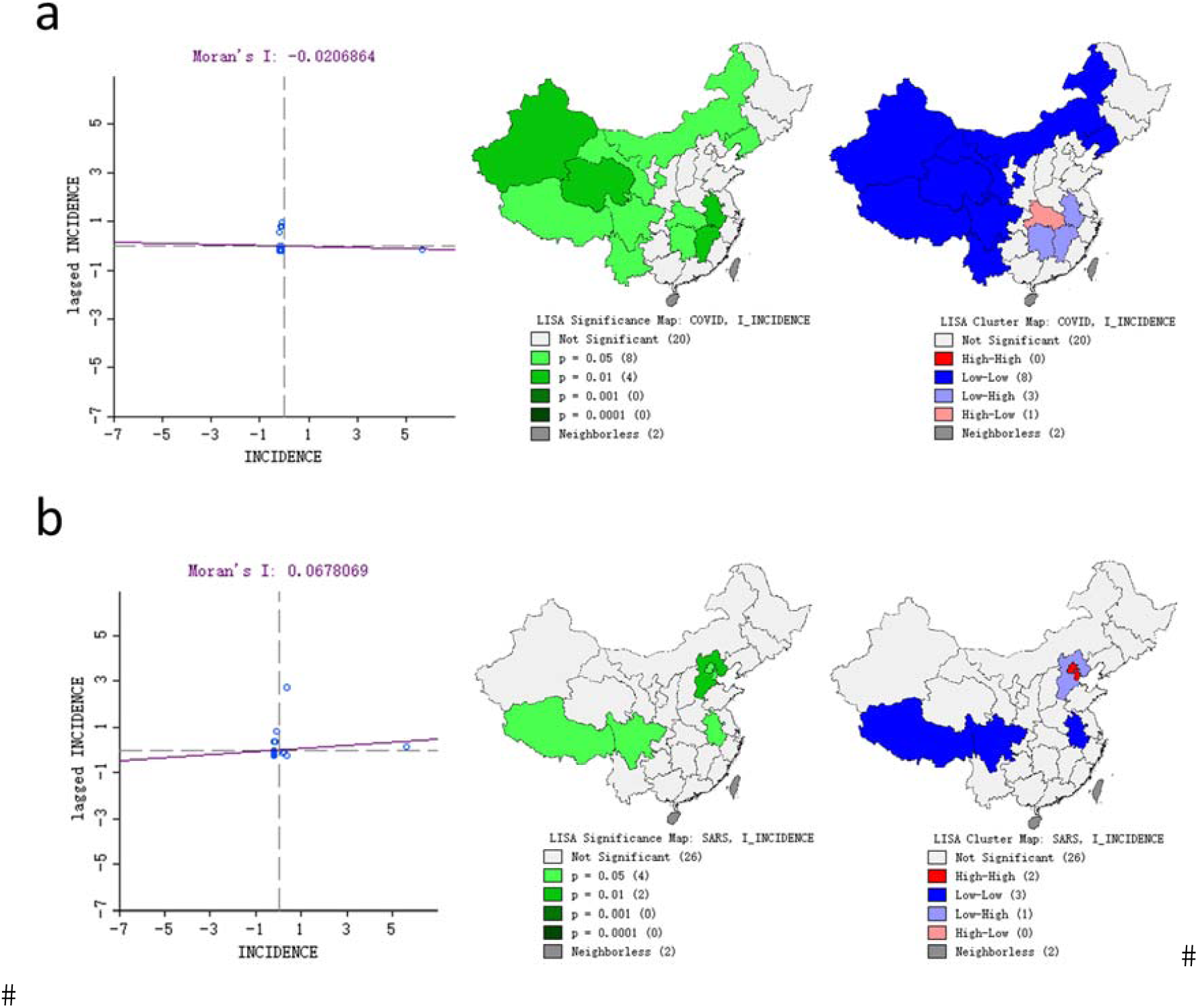
Moran scatter plot and LISA cluster map for COVID-19 (a) and SARS (b).

The LISA cluster map showed the significant locations color coded by the type of spatial autocorrelation. For COVID-19, the high-low spatial clustering was in Hubei Province. In addition, we identified 4 significant clusters at *P* < 0.01 and 5 significant clusters at *P* < 0.05. Specifically, Liaoning, Inner Mongolia, and most western provinces had significantly low-low spatial clustering, whereas Anhui, Hunan and Jiangxi of Central China had significantly low-high spatial clustering. For SARS, two significant high-high (Beijing and Tianjin) and low-high (Hebei) clusters were detected. Sichuan, Tibet and Anhui showed significant low-low clustering (**Fig. 4**).

## Discussion

In our study, we found that there were different spatiotemporal clustering patterns between COVID-19 and SARS. Compared to SARS, COVID-19 had a higher incidence as well as wider and faster transmission in mainland China. The significant high-risk areas for COVID-19 were predominantly clustered in south-central China, around Hubei, from Jan. 28 to Feb. 18. Additionally, our results showed that the quarantine measure taken in Hubei might have played a crucial role in restricting the infected areas, shortening the epidemic period, and reducing the national infected risk of the disease.

The 2003 SARS outbreaks represented one of the most serious public health challenges to China and the world [12]. Seventeen years later, the outbreak of COVID-19 could encounter a similar situation but lead to a different outcome. The different transmission mechanisms of these coronaviruses can also present different spatial and temporal distributions nationally and globally. For SARS, we observed that the distance transmission chain started from Guangdong to Beijing and the nearby provinces. However, for COVID-19, we observed a shorter transmission chain around Hubei but a wider infected region nationally. Outside the epicenter, we identified more secondary clusters for SARS, which indicated that the transmission was wider for second generations. Compared to SARS, the secondary clusters of COVID-19 were mainly clustered around Hubei. This could be explained by the relatively high infection rate nationally, as well as the different demographic factors and local government containment strategies regionally. Another secondary cluster identified in Shandong around Feb. 20 was mainly due to the newly reported cases previously identified in jails [13]. Because the reporting system of the jails was independent from the national reporting system, these cases were reported with a time lag. When these cases were not considered, the incidence was generally low during the last two weeks in February.

Because the mandatory quarantine for Hubei (“Wuhan lockdown”) has been in effect since Jan. 23, 2020, and because social-distancing measures, such as population movement restrictions, school closures and temperature monitoring at public locations, have also been in effect in most provinces in mainland China since this date, we distinguished the spatial patterns of the COVID-19 epidemic before and after this date plus a 14-day incubation period. We found that COVID-19 cases were clustered mainly in Hubei, and other secondary clusters disappeared, except in Shandong. This reinforced that quarantine and isolation can help to contain the virus, prevent transmission and effectively reduce the number of secondary clusters.

The present study was based on the daily briefings provided by the health department. Although our study covered the peak period of the outbreaks in most provinces, transmission patterns of SARS in Beijing and Guangdong were biased due to lack of the available data before Apr. 21, 2003. Comparatively, reported cases for COVID-19 could also be biased due to missing data before Jan. 21, 2020 in this study. Moreover, early infections with atypical presentations may have been missed [14]. Therefore, the conclusions should be interpreted with caution with regard to the early stages of the two epidemic outbreaks. When more data become available on transmission patterns and epidemiologic characteristics of COVID-19 and SARS, a detailed comparison with the corresponding characteristics between the two diseases would be more informative.

## Data Availability

NA.

## Acknowledgements

We would like to show our respect and gratitude to all the health workers who are at the front line of the outbreak response and fighting against COVID-19 in China.

## Funding

There was no funding source for this research.

## Conflict of interest

The authors declare that they have no conflicts of interest.

## Authors’ contributions

XZ and HD conceived and designed the study. XZ, RQ, YW and HD collected the data. XZ and RQ performed the statistical analysis. XZ and HD prepared the manuscript. All authors read and approved the final manuscript.

## Figure legends

**Supplementary Figure 1**. Spatiotemporal clustering of COVID-19 incident cases in stage1 (a) and stage2 (b) (including Hubei).

**Supplementary Figure 2**. Spatiotemporal clustering of COVID-19 incident cases in stage1 (a) and stage2 (b) (excluding Hubei).

## Notes

### Competing Interest Statement

The authors have declared no competing interest.

